# Social Network Strategies to Distribute HIV Self-testing Kits: A Global Systematic Review and Network Meta-analysis

**DOI:** 10.1101/2023.11.05.23298135

**Authors:** Siyue Hu, Fengshi Jing, Chengxin Fan, Yifan Dai, Yewei Xie, Yi Zhou, Hang Lv, Xi He, Dan Wu, Joseph D. Tucker, Weiming Tang

## Abstract

**Introduction:** Social network strategies, in which social networks are utilized to influence individuals or communities, are increasingly being used to deliver human immunodeficiency virus (HIV) interventions to key populations. We summarized and critically assessed existing research on the effectiveness of social network strategies in promoting HIV self-testing (HIVST).

**Methods:** Using search terms related to social network interventions and HIVST, we searched five databases for trials published between January 1^st^, 2010, and June 30^th^, 2023. Outcomes included uptake of HIV testing, HIV seroconversion, and linkage to antiretroviral therapy (ART) or HIV Care. We used network meta-analysis to assess the uptake of HIV testing through social network strategies compared with control methods. A pairwise meta-analysis of studies with a comparison arm that reported outcomes was performed to assess relative risks (RR) and their corresponding 95% confidence intervals (CI).

**Results and discussion:** Among the 3,745 manuscripts identified, 33 studies fulfilled the inclusion criteria, including one quasi-experimental study, 17 RCTs and 15 observational studies. Networks HIVST testing was organized by peers (distributed to known peers, 15 studies), partners (distributed to their sexual partners, 10 studies), and peer educators (distributed to unknown peers, 8 studies). The results showed that all of the three social network distribution strategies enhanced the uptake of HIV testing compared to standard facility-based testing. Among social networks, peer distribution had the highest uptake of HIV testing (79% probability, SUCRA 0.92), followed by partner distribution (72% probability, SUCRA 0.71), and peer educator distribution (66% probability, SUCRA 0.29). Pairwise meta-analysis showed that peer distribution (RR 2.29, 95% CI 1.54-3.39, 5 studies) and partner distribution (RR 1.45, 95% CI 1.05-2.02, 7 studies) also increased the probability of detecting HIV reactivity during testing within the key population when compared to the control. Linkage to ART or HIV Care remained comparable to facility-based testing across the three HIVST distribution strategies.

**Conclusions:** Network-based HIVST distribution is considered effective in augmenting HIV testing rates and reaching marginalized populations compared to facility-based testing. These strategies can be integrated with the existing HIV care services, to fill the testing gap among key populations globally.

PROSPERO Number: CRD42022361782

## Introduction

Key populations, such as men who have sex with men (MSM), people who use drugs, and sex workers, face a heightened risk of HIV exposure [1]. The contribution to HIV transmission is greater than that of low-risk populations outside this population [2, 3]. Accordingly, the public health community increasingly recognizes that key populations have a unique role in implementing HIV interventions. As a complementary approach to HIV testing services (HTS), HIV self-testing (HIVST) has demonstrated its safety, accuracy, and acceptability among key populations [4, 5]. With HIVST, individuals can collect their samples for testing and read the results to decide where and when to test while ensuring efficiency, privacy, and confidentiality [6]. The significance of HIVST is underscored in the latest consolidated guidelines on HIV, while the WHO has further emphasized recommending network-based strategies in their updated recommendations [7, 8]. Thus, strategies that can facilitate the effectiveness of HIVST are needed to achieve the maximum prevention effect of HIVST.

Social network strategy, a method or approach used in social interventions that operates through interconnections among at-risk groups, has great potential to improve HIVST coverage [9]. In the social network context, peers share the same characteristics (e.g., demographic, cultural, health outcomes, and behaviors) with the target audience [10]. Members of social networks often have similar HIV risks, trust each other, and are interested in helping each other [11]. Based on these principles, our social network strategies utilize peer-driven interventions in which peers (with or without prior knowledge) take on the role of HIV prevention educators, HIVST kit providers, or health partners to help identify network partners and motivate others to get tested. Information can be disseminated more effectively when health information and incentives for preventive measures are disseminated through close social relationships [12]. Using close friends or trusted peers as a source of information increases the credibility and trustworthiness of the information for the recipients [13]. In addition, social network strategies utilize social pressure and reciprocal relationships to encourage individuals to adopt healthy behaviors and provide support when needed [14, 15]. Social network strategies aim to harness social networks to generate social influence and accelerate behavior change to achieve desired outcomes at the individual or community level. HIV testing based on social network strategies has been supported by CDC, WHO, and other guidelines while increasingly being used in HIV interventions [8, 16].

Previous studies have mainly evaluated the efficiency of HIVST, which have shown higher test uptake rates compared to standard testing services alone [5]. Meanwhile, studies that have explored the role of social networks have generally only compared HIV testing or risk behaviors and have not further explored the impact of social networks on HIVST [14, 17]. Some of these studies have been limited to marginalized populations such as female sex workers [18]. Currently, conversations around HIV testing have shifted towards integrating HIVST as a strategy within the continuum of HIV care, and social network strategies have shown great potential in improving HIVST [15, 19], while there is an expansion of network-supported HIVST. However, there is a lack of reviews that summarize the role of social network strategies in HIVST interventions. Knowledge gaps remain in terms of which strategies are most effective, for whom, and in which settings they are best suited for scaling up. To optimize the effectiveness of HIVST, it is essential to consider information about the role of social networks in the HIV care cascade. Network meta-analysis allows for simultaneous comparison of the effects of multiple interventions and consideration of other potential sources of heterogeneity. Using direct comparisons to generate indirect effect estimates and ranking distribution strategies provides a complementary approach to determining optimal implementation strategies.

This study aims to investigate the effects of social network strategy on improving HIV testing uptake among at-risk populations who used HIVST through a systematic review and network meta-analysis.

## Methods

### Search Strategy and Selection Criteria

This systematic review and meta-analysis followed the Preferred Reporting Items for Systematic Reviews and Meta-Analyses for Network Meta-Analyses (PRISMA-NMA) and Cochrane guidelines [20]. The following databases were utilized for the literature search: (1) PubMed; (2) Embase; (3) Web of Science; (4) Cochrane Library; and (5) Wiley. The initial search strategy formulated for PubMed used the combination of key terms that include “HIV/AIDS”, “social network”, and “HIVST” (Supplementary Appendix 1), which were subsequently adapted into corresponding index terms for the other searched databases. The search was completed and limited to peer-reviewed journal articles published in English from January 1st, 2010, to June 30th, 2023, with no geographic limitation. Additional articles were identified through manual reference checking of relevant studies.

Trials with a comparison arm or observational studies that evaluated any social network strategies used for HIVST in any setting that reported quantitative outcomes were included. The study population was the population receiving HIVST services. All studies were required to use social network strategies as an intervention and to report outcomes on HIV testing uptake, HIV seroconversion rates, or linkage to ART or HIV Care. ART initiation was selected preferentially as an outcome, and linkage to any HIV services was used if ART initiation was not available. Studies without a comparison arm were also included. Two independent reviewers (YD and CF) first assessed the title and abstracts to identify relevant records for inclusion following the eligibility criteria, with a third reviewer as a tiebreaker (SH). Full texts of included studies were retrieved and assessed for inclusion following the same screening method. Two reviewers jointly developed a data extraction form and completed it independently.

### Intervention categorization

We grouped social network strategies for HIVST according to who distributed the tests, yielding three distribution strategies: 1) peer distribution focused on recruiting peers (defined as “index”) and encouraging peers to distribute the HIVST kits to people in their social networks (known peers, defined as “alter”); 2) partner distribution that distributed the kits from participants to their sexual partners; and 3) peer educator distribution that peer educators distribute the services to people outside their social network (unknown peers). Peer educators were usually assigned to recruit participants or were randomly assigned to groups of recruited participants after receiving corresponding training, and then linkages were established for behavioral interventions. Peer educators acted as HIV-related information popularizers and HIVST kit providers to influence social norms in the established community for further distribution of the HIVST kits.

### Data analysis

For studies with a comparison arm, pooled relative risks (RR) with 95% confidence intervals (CI) were used to compare outcomes and the heterogeneity was assessed by calculating I2. We built forest plots for each outcome using Review Manager version 5.4. A network meta-analysis was performed to analyze the primary outcome of HIV testing uptake. The final model was selected by evaluating a combination of the deviance information criterion (DIC), Markov chain Monte Carlo (MCMC) error, and trace and density plots. The node-splitting model was applied to analyze the direct and indirect comparison results to observe the consistency. Results were presented in risk ratios (RR) and 95% credible intervals (CrI), as well as relative effects tables and forest plots. In addition, ranking probabilities were used to indicate the likelihood or chance of each intervention being ranked at a specific position within the comparison. A ranking probability of 1 (100%) represents the highest ranking of a distribution strategy and 0 the lowest. Probability values were summarized and are reported as the surface under the cumulative ranking (SUCRA). All network meta-analyses were carried out using the meta and gemtc packages in R software version 4.2.2.

### Quality assessment

In an analysis of quality assessment, studies were stratified based on study design and level of evidence. Bias among randomized controlled studies were assessed across five domains using the Cochrane risk of bias tool [21]. Bias in observational studies was assessed using the Newcastle-Ottawa Quality Assessment Scale [22]. The quality of observational studies was evenly distributed, with some studies judged to be of poor or fair quality, mainly due to insufficient comparability and outcome data (Supplementary Table 5). For all randomized controlled trials (RCTs), performance and detection bias items represented a high risk of bias, respectively, primarily due to failure to blind participants and personnel and to blind outcome assessment (Supplementary Table 6 and Supplementary Figure 1). A quasi-experimental study identified concerns about selection and reporting bias due to selective reporting.

## Results

The search yielded 3,640 citations and 105 additional records identified through other sources. After removing duplicate citations, 2,775 unique titles and abstracts were screened, and 73 full-text articles were assessed for eligibility. This evaluation ultimately included 33 studies, including one quasi-experimental study, 17 RCTs, and 15 observational studies (Figure 1). Six studies were from high-income countries, 19 in middle-income countries, and eight in low-income countries. The included studies were conducted in eight countries from 2010-2021, six in China [23–28], six in South Africa [29–34], six in the US [35–40], five in Uganda [41–45], four in Kenya [46–49], three in Malawi [50–52], two in Vietnam [53, 54], and one in Zambia [55].

**Figure 1.**
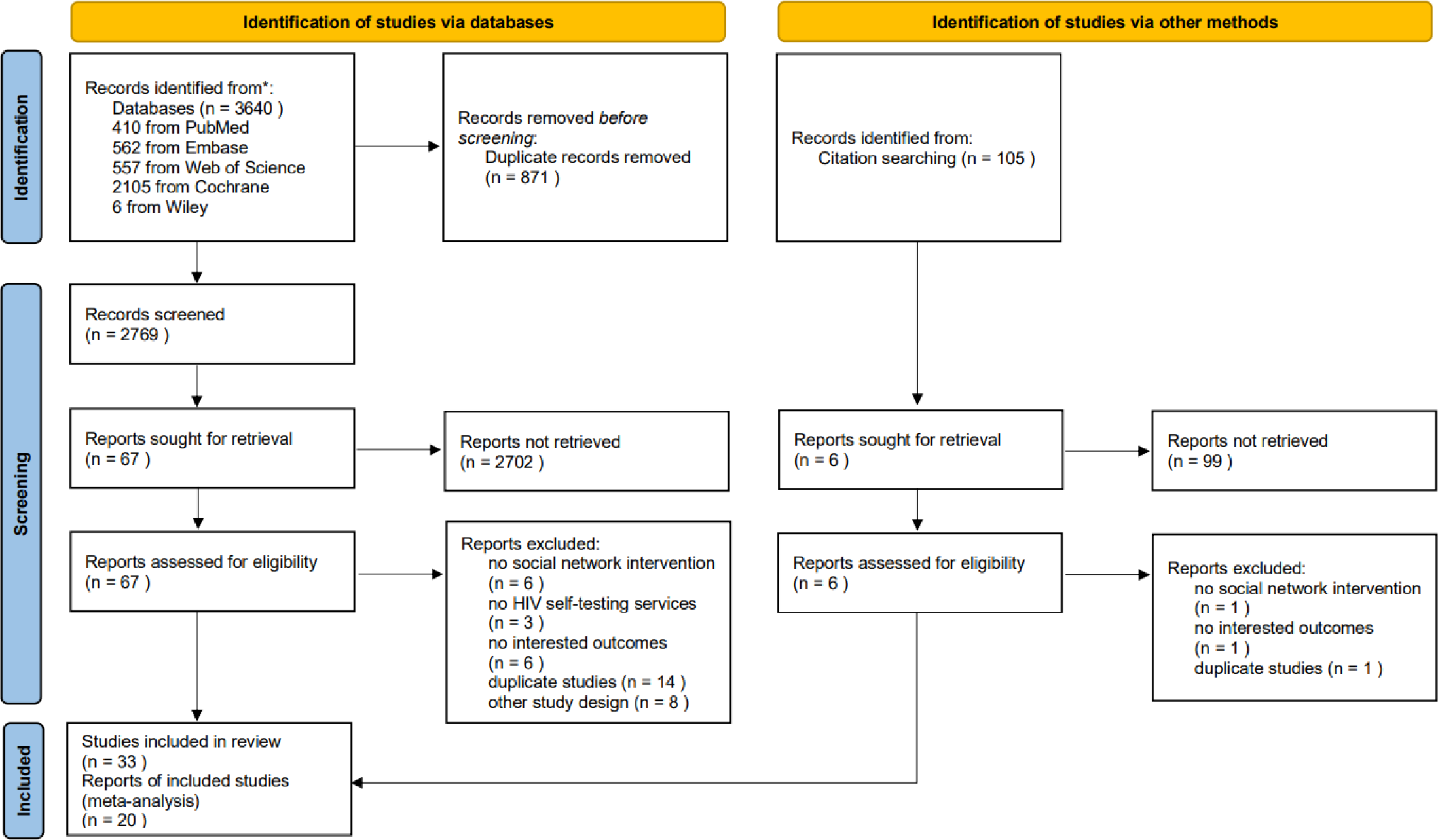
PRISMA-NMA flow chart for study.

The study populations were: 15 conducted in MSM, three in male partners of antenatal care clinic (ANC), two in key populations and their partners (including people who inject drugs (PWID), MSM, sex workers, and their partners), two in female sex workers (FSW), two in young people, two in young woman and their partners, two in women living with HIV and their male partners, one in partners of people living with HIV, one in general population and three in two populations. Among them, two were conducted in partners of people living with HIV and male partners in ANC, and one in both key populations and sexual partners of pregnant and lactating women.

Of these included studies, 21 of the 33 studies had comparison groups, 17 studies compared HIVST distribution with standard HCW-administered facility-based HIV testing, one study compared different forms of the secondary distribution of HIVST, and three studies compared the impact of peer educators on HIVST distribution. Regarding compensation, 16 studies had no compensation, whereas 12 studies paid their partners, with 11 giving monetary incentives and one giving material incentives, including reimbursement for transportation, T-shirts, backpacks, and umbrellas. In addition, five studies imposed additional requirements, of which two had to advise on partner negotiation and communication, one required telephone confirmation that the kit had been delivered to the recipient, one required a report for confirmatory HTC at the clinic, and one required daily or on-demand PrEP. The characteristics of the included studies and interventions with and without comparison groups are shown in Table 1A and Table 1B. Further detailed characterization and study outcomes are listed in the Supplementary Material (Supplementary Tables 1-2).

**Table 1A.**
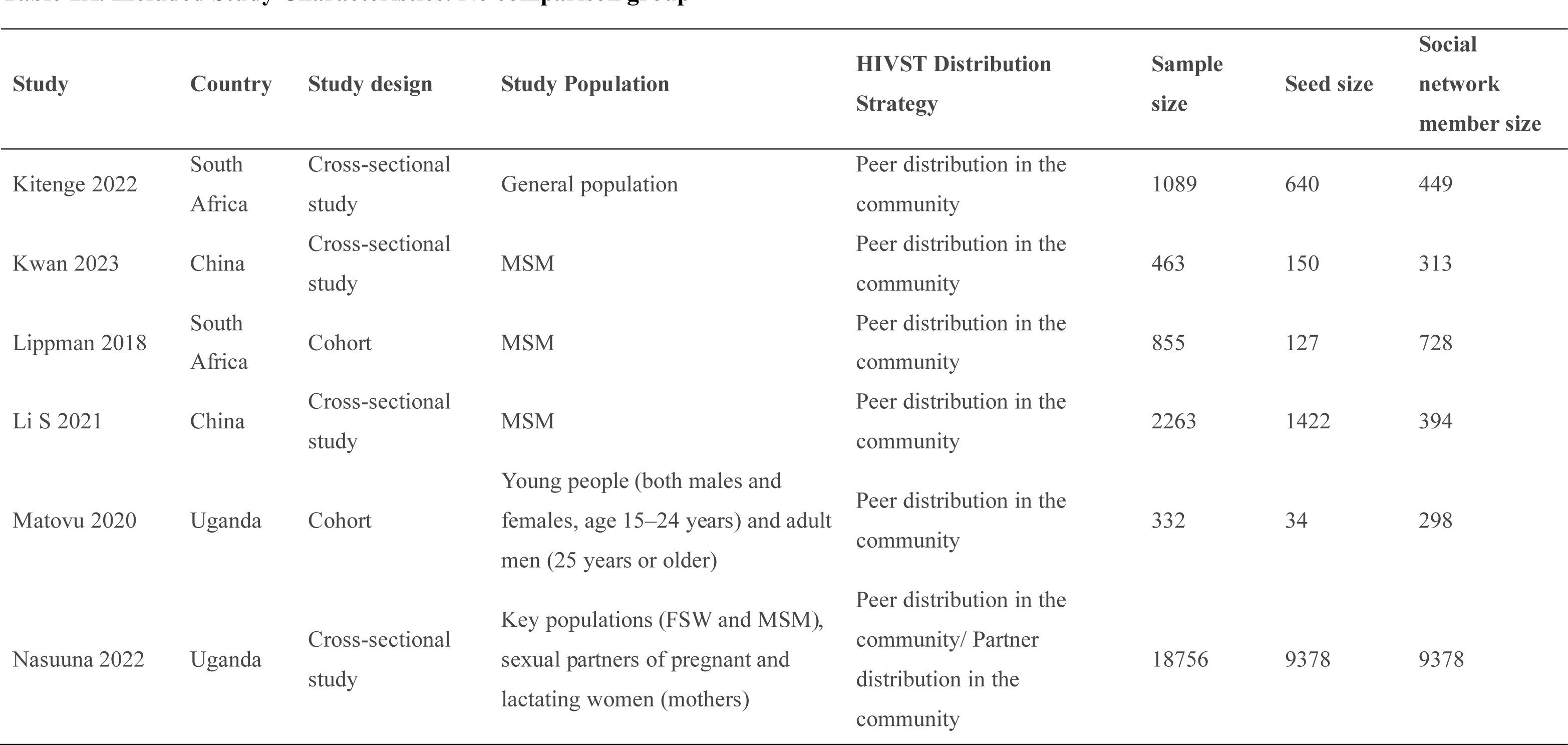
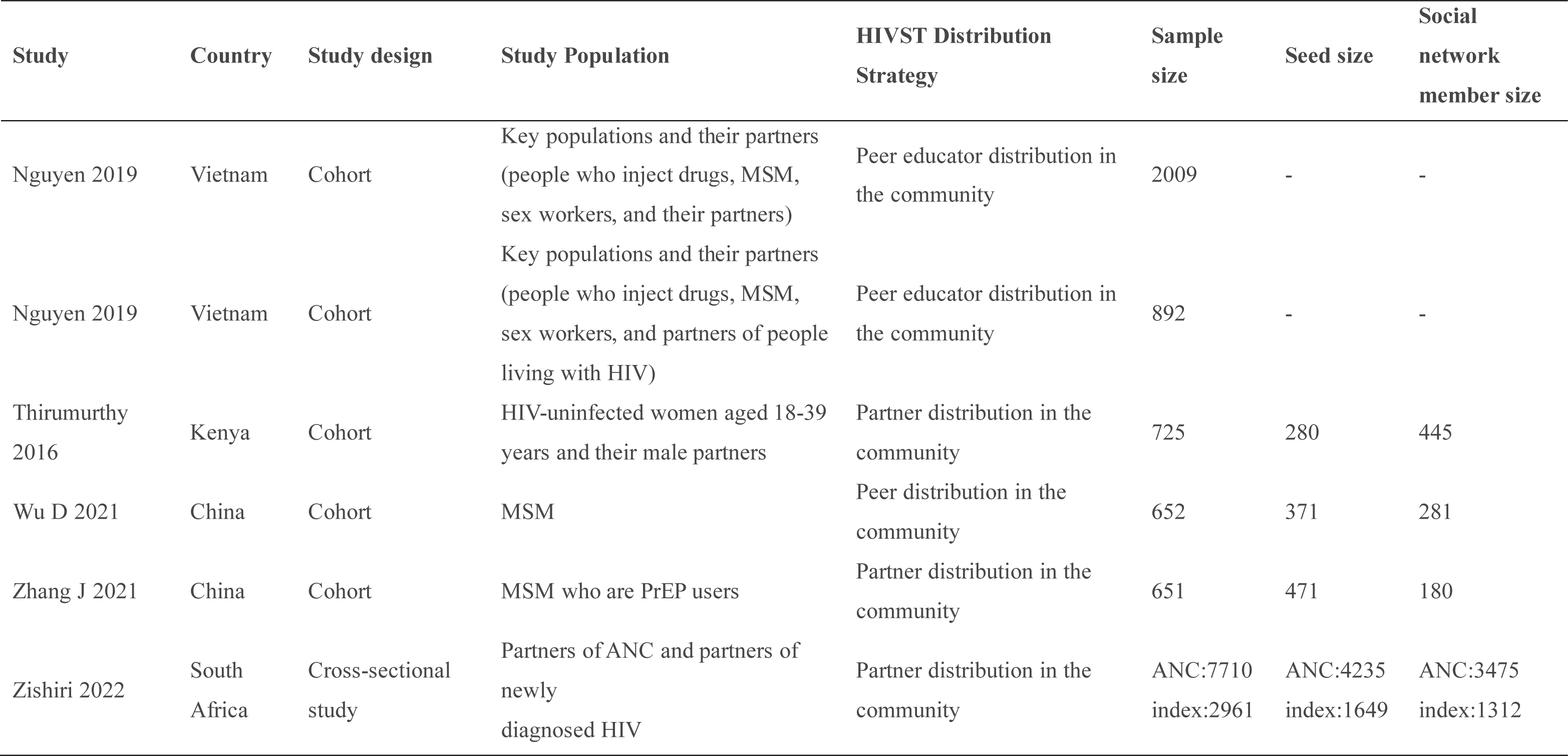
Included Study Characteristics: No comparison group.

**Table 1B.** Included Study Characteristics: Comparison group.

| Study | Country | Study design | Study Population | HIVST Distribution Strategy | Sample size | Seed size | Social network member size |
| --- | --- | --- | --- | --- | --- | --- | --- |
| Chanda 2017 | Zambia | Cluster RCT | FSW | Peer educator distribution in the community | 1125 | 160(Standard:53, Delivery: 53, Coupon: 54) | 965(Standard:320, Delivery: 316, Coupon: 329) |
| Choko 2019 | Malawi | Cluster RCT | Male partners of ANC | Partner distribution in the community | 4698 | 2349(I:1941, C:408) | 2349(I:1941, C:408) |
| Choko 2021 | Malawi | Cluster RCT | Partners of ANC, partners of newly diagnosed HIV | Partner distribution in the community | 8505 | ANC:4544(SOC:1447, HIVST:1465, HIVST plus:1632) index:708(SOC:234, HIVST:169, HIVST plus:305) | ANC:2604 (SOC:498, HIVST: 1106, HIVST plus:1000) index:649 (SOC:209, HIVST:155, HIVST plus:285) |
| Dovel 2019 | Malawi | RCT | Partners of people living with HIV | Partner distribution in the community | 849 | 484(I:349, C:135) | 365((I:258, C:107) |
| Frye 2021 | The US. | RCT | YBMSM (young black MSM) and Transwomen | HCW at the health facility | 376 | 188(I:89, C:99) | 188(I:90, C:98) |
| Gichangi 2018 | Kenya | RCT | Male partners of ANC | Partner distribution in the community | 2543 | 1410(I:472, C:938) | 1133(I:396, C:737) |
| Joseph 2022 | South Africa | RCT | Male partners of WLHIV (women living with HIV) | Partner distribution in the community | 352 | 176(I:85, C:91) | 176(I:85, C:91) |
| Lightfoot 2018 | The US. | Cohort | MSM | Peer distribution in the community | 195 | 30 | 165 |
| MacGowan 2020 | The US. | RCT | MSM | Peer distribution in the community | 2968 | 667 | 2301 |
| Masters 2016 | Kenya | RCT | Male partners of ANC | Partner distribution in the community | 1140 | 570(I:284, C:286) | 570(I:284, C:286) |
| Merchant 2018 | The US. | RCT | YMSM (young adult black, Hispanic, and white MSM) | Peer distribution in the community | 241 | 88(I:51, C:37) | 153(I:82, C:71) |
| Mujugira 2022 | Uganda | RCT | Male partners of PWLHIV (pregnant women living with HIV) | Partner distribution in the community | 723 | 489(I:328, C:161) | 234(I:159, C:75) |
| Okoboi 2020 | Uganda | Cross-sectional study | MSM | Peer distribution in the community | 165 | 15 | 150 |
| Ortblad 2017 | Uganda | Cluster RCT | FSW | Peer educator distribution in the community | 960 | 120(Direct: 37, Facility: 42, Standard: 41) | 840(Direct: 259, Facility: 294, Standard: 287) |
| Pettifor 2020 | South Africa | RCT | Young women and their peers and partners | Peer distribution in the community | 1394 | 287(I:141, C:146) | 1107(I:701, C:406) |
| Sha 2022 | China | Quasi-experimental study | GBMSM (gay, bisexual, and other MSM) | Peer distribution in the community | 359 | 154(I:92, C:62) | 205(I:179, C:26) |
| Shahmanesh 2021 | South Africa | Cluster RCT | Young women (18–24) and all young people (aged 18–30) | Peer distribution in the community | 4220 | 57(I: 38, C: 19) | 4163(I: 3065, C: 1098) |
| Young 2013 | The US. | Cluster RCT | MSM | Peer educator distribution in the community | 128 | 16(I:8, C:8) | 112(I:55, C:57) |
| Young 2022 | The US. | RCT | MSM | Peer educator distribution in the community | 979 | 79(I:79, C:0) | 900((I:450, C:450) |
| Van Der Elst 2017 | Kenya | Cohort | MSM | Peer distribution in the community | 1038 | 11(I:6, C:5) | 1027(I:337, C:690) |
| Zhou 2022 | China | RCT | MSM | Peer distribution in the community | 653 | 309(C:102, SD-M:103, SD-M-PR:104) | 344(C:58, SD-M:101, SD-M-R:185) |

### HIVST Distribution Strategy

We classified the social network interventions into three categories based on the people who distributed the test: peers, partners, and peer educators. Twenty-five studies used risk-based framing for caring about peers and partners for their at-risk or disease. Eight studies used asset-based framing because peer educators have strengths and insight.

Overall, 15 studies recruited index participants and encouraged participants to distribute them to people in their social networks (alters) [23–25, 27, 28, 30, 32, 33, 35, 39, 40, 43–45, 48]. Most studies had multiple HIVST kits provided directly to index participants by healthcare workers (HCWs) for distribution to alters. Two studies used formative research by conducting focus group discussions to collect the necessary data to inform the design of peer-led HIVST interventions [39, 44]. Specifically, the peer training focused on basic counseling, approaching social network members, using HIVST kits, and linkage to HIV services. Ten studies distributed HIVST kits directly from participants to their sexual partners [26, 31, 34, 42, 46, 47, 49–52].

Two of these studies additionally provided participants with techniques for negotiating and communicating with their sexual partners about using HIVST kits, and the other two studies explored the impact of financial incentives [50, 52]. Peer educators were recruited in eight studies to provide additional interventions [29, 36–38, 41, 53–55]. The peer educators received training varying in length from 9h-20 weeks, divided into how to provide pre-test and post-test counseling to participants. Pre-test counseling focuses on basic counseling, communication skills, and how to use HIVST. Post-test counseling focuses on interpreting test results, referral skills, and linkage to HIV care services [41, 53–55].

### Role of Social Network

Social networks played more than one role in these 33 studies. The relatively small size of the at-risk population and the privacy issues make it difficult to reach through routine sampling.

Participants distributed HIVST kits within their social circles, and social networks were used to access hard-to-reach individuals to facilitate more comprehensive coverage of potential users who do not typically use the health system [23, 30, 31, 35, 42, 45–47, 50–52]. Secondary distribution of HIVST kits for pregnant women has been more effective than other methods in increasing HIV testing, and it has shown great potential for reaching sexual contacts of newly diagnosed HIV [50, 52].

As a platform for providing services such as HIVST kits, HIV self-sampling, or home-based HIV testing, social networks also provide a better channel for communication and diffusion. Through this channel, not only can HIV testing be offered to key populations, but it can also provide a platform for communicating HIV risk information while collecting data information on testers in the process. By recruiting index participants, participants are encouraged to distribute HIVST kits to members in their social networks face-to-face or by sharing online links and mail. In most studies, HIVST kits were distributed person-to-person, with participants applying for HIVST kits and then distributing them face-to-face in their social networks [23, 26, 30–34, 42, 46, 47, 49–52]. In contrast, a subset of studies were based on social media, an HIVST application link on a phone app [24, 25, 27], or a web-based platform [28, 35, 40] that provides HIVST kits in the form of online ordering and mailing.

### Uptake of HIV testing

Data on the uptake of HIV testing were obtained from 16 studies. Network meta-analysis directly compared differences in the uptake of HIV testing in HIVST kits distributed by peers (4 studies), partners (8 studies), and peer educators (4 studies) in their social networks with the standard HCW-administered facility-based HTS (Figure 2). Network estimates showed that the distribution of peer in social network (RR 2.60, 95% CrI: 1.40-5.30) and partner in sexual network (RR 2.00, 95% CrI: 1.23-2.44) methods resulted in higher HIV testing uptake than facility-based testing (Figure 3). However, the Effect of peer educator distribution on the increased HIV testing uptake was insignificant (RR 1.20, 95% CrI: 0.68-2.22). Compared with facility-based (76% of simulations with the last rank, SUCRA 0.08), ranking probabilities (Figure 4) showed that HIVST kits had the highest uptake among social networks via peer distribution (79% of simulations with the highest rank, SUCRA 0.92), partner distribution (72% of simulations with the second highest rank, SUCRA 0.71) and peer educator distribution (66% of simulations with the third highest rank, SUCRA 0.29) (Supplementary Table 3).

**Figure 2.**
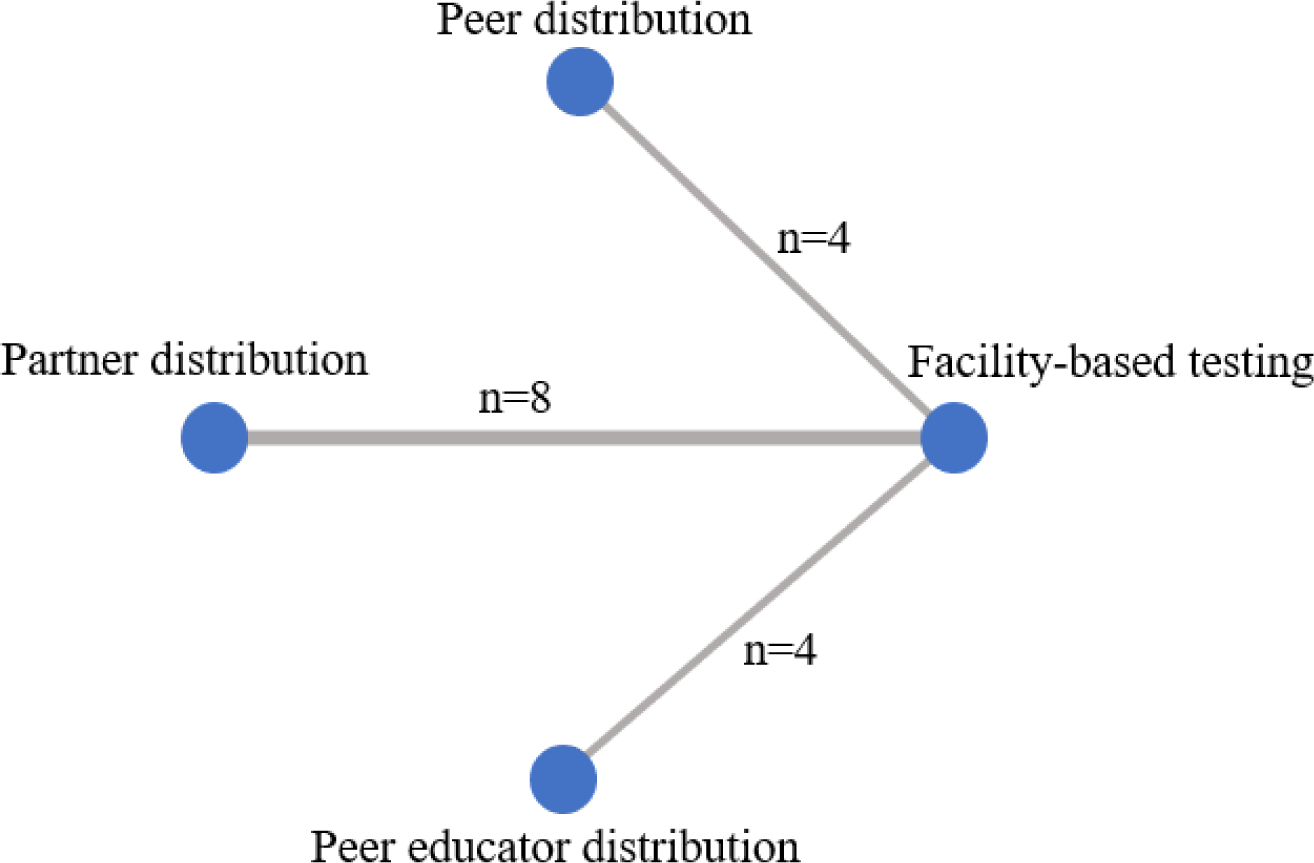
Network map: uptake of HIV testing. Network map represents the number of studies contributing to the direct comparisons in the network.

**Figure 3.**
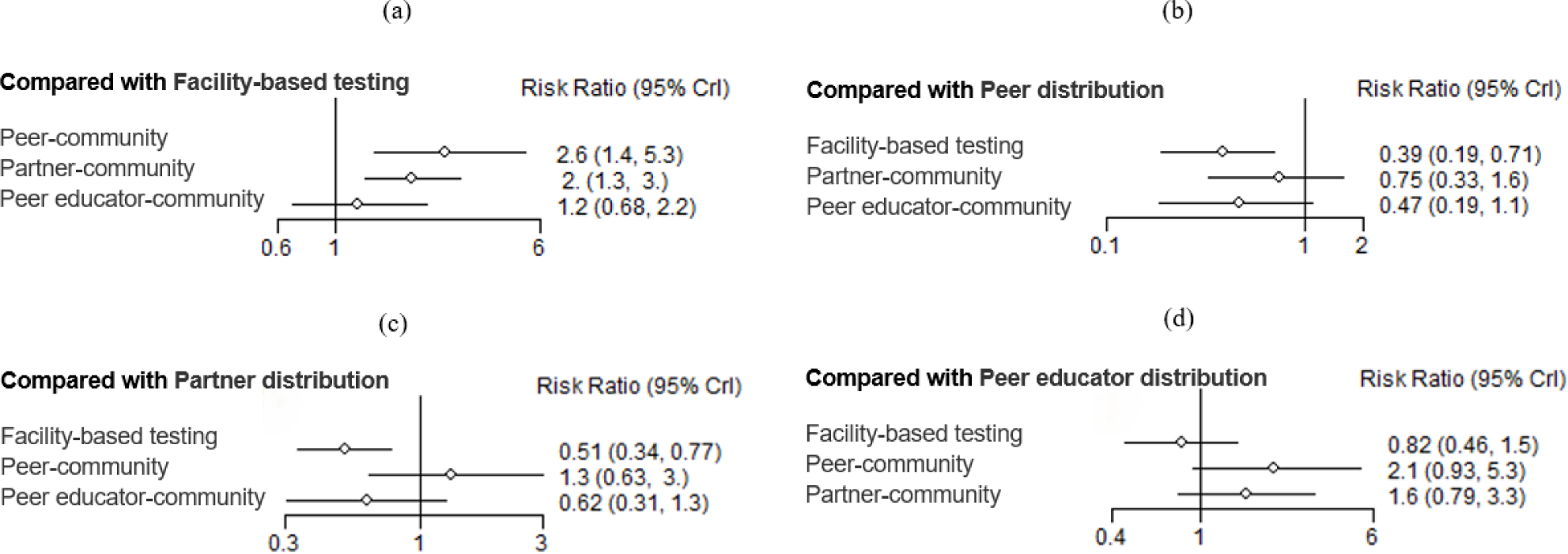
Network estimates of HIV testing uptake.

**Figure 4.**
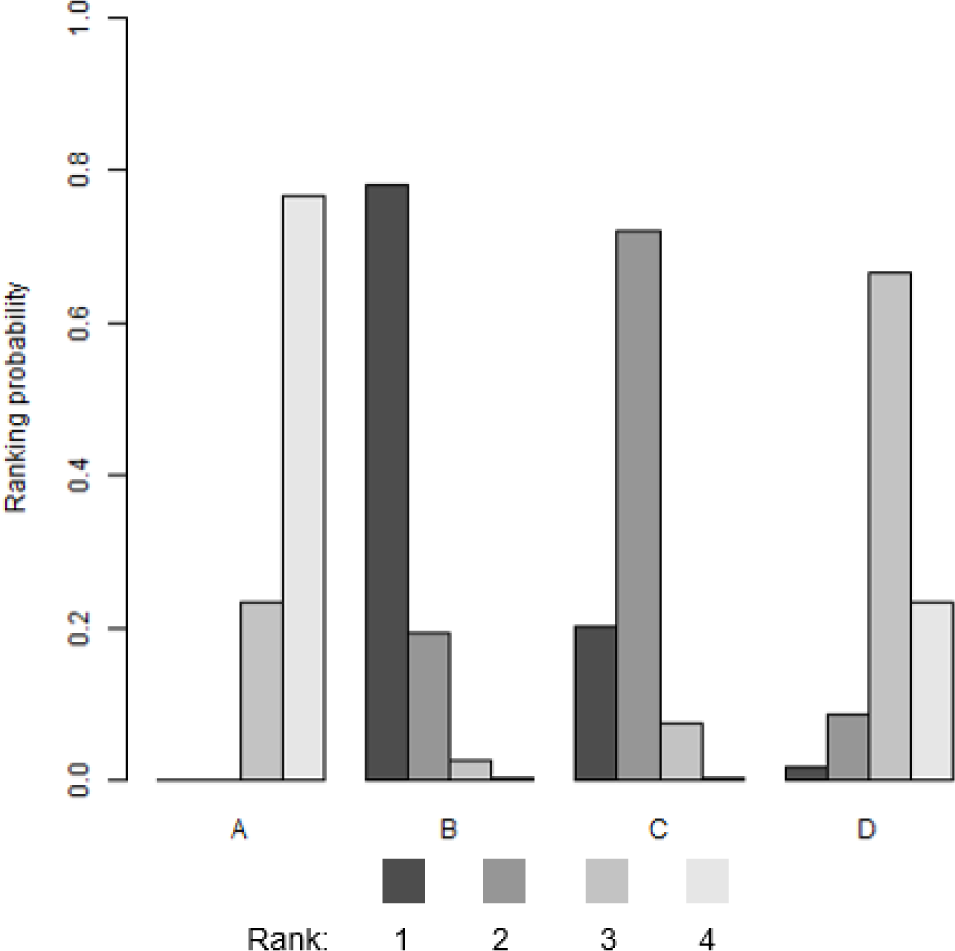
HIV testing strategies ranking probabilities for HIV testing uptake. For each strategy, the colored bars represent the probability that that strategy ranks first, second, third, and so forth. Darker colors represent high ranking (most effective); light colors represent low ranking (least effective). A: Facility-based testing, B: Peer distribution, C: Partner distribution, D: Peer educator distribution.

Data from pairwise meta-analyses also supported this view. Four studies reported uptake of HIV testing through peer distribution [23, 30, 35, 39]. The meta-analysis showed a doubling of HIV testing uptake compared with standard of care (SOC), the standard facility-based testing (RR 2.10, 95% CI 1.42-3.10, I2 66%). Data from seven RCTs showed that HIV testing rates were significantly higher when social network interventions were delivered through sexual partners than in the SOC (RR 1.95, 95% CI 1.24-3.06, I2 99%) [31, 42, 46, 47, 50–52]. Similarly, findings showed that the intervention group that used social influence with the help of peer educators to disseminate HIV risk information and attempted to change social norms with the help of peer educators (RR1.18, 95% CI 1.12, 1.25], I2 48%) had higher uptake of HIV testing than the comparison group (Figure 5) [36, 38, 41, 55].

**Figure 5.**
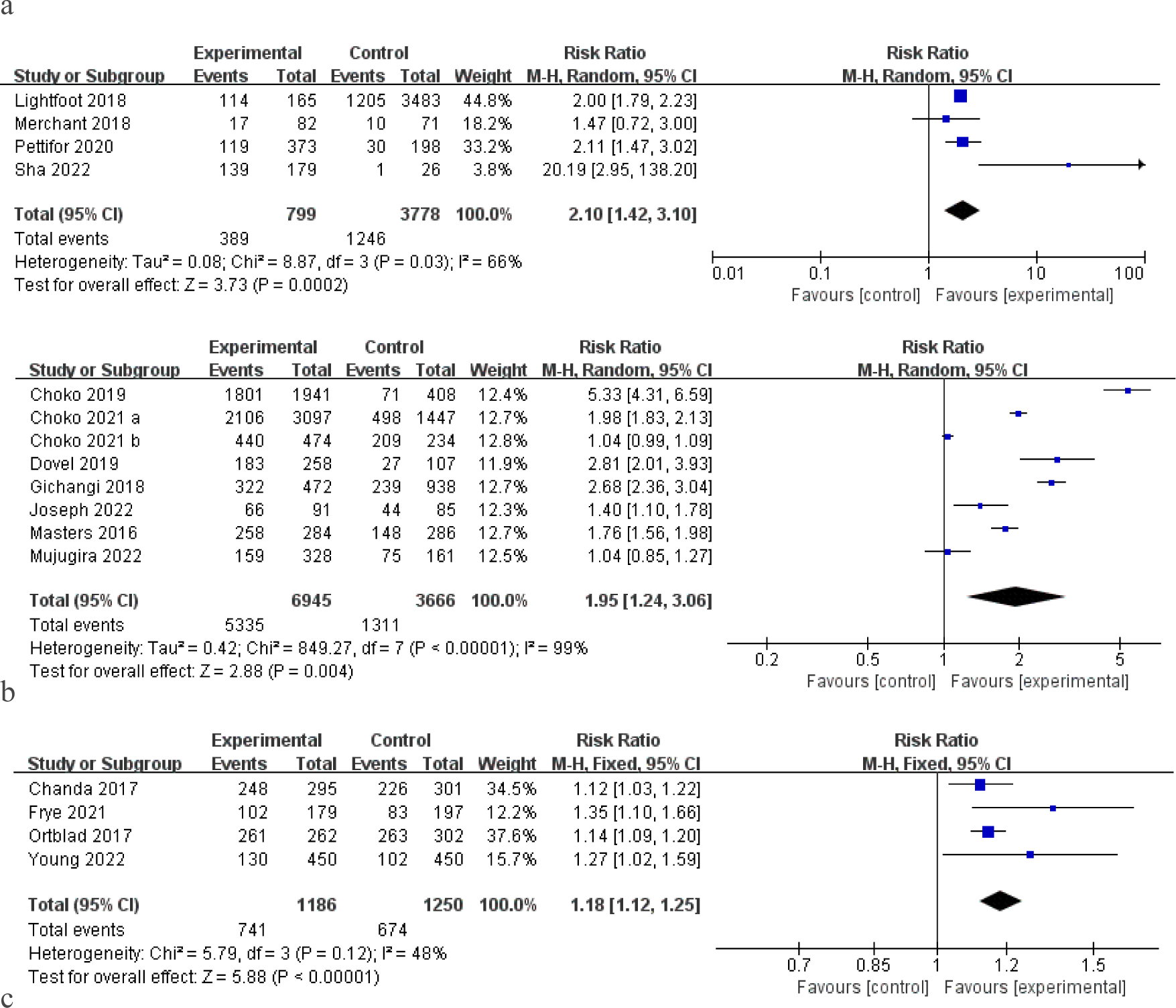
Uptake of HIV testing by distribution strategy. figure 5(a): Peer distribution in community, figure 5(b): Partner distribution in community, figure 5(c): Peer educator distribution in community.

### HIV seroconversion

Overall, 13 studies reported HIV seroconversion after HIV testing by comparing HIV reaction rates among alters in social networks who received HIVST kits. Five studies that reported differences in HIV seroconversion outcomes based on peer distribution were included [23, 30, 39, 43, 48]. Meta-analysis showed significantly higher HIV reaction rates among alters using peer distribution compared to SOC (RR 2.29, 95% CI 1. 54-3.39, I2 52%). Data on HIV reaction rates for partner distribution were obtained from seven studies [31, 42, 47, 50–52]. Interventions using partner-distributed HIVST kits showed a higher likelihood of detecting HIV reactivity in the partners of participants (RR 1.45, 95% CI 1.05-2.02, I2 5%). However, when comparing the HIVST distribution strategy using peer educator influence to the comparison group, there appeared to be no difference in the association of finding people living with HIV (RR 0.91, 95% CI 0. 74-1.13, I2 0%) (Figure 6) [41, 55].

**Figure 6.**
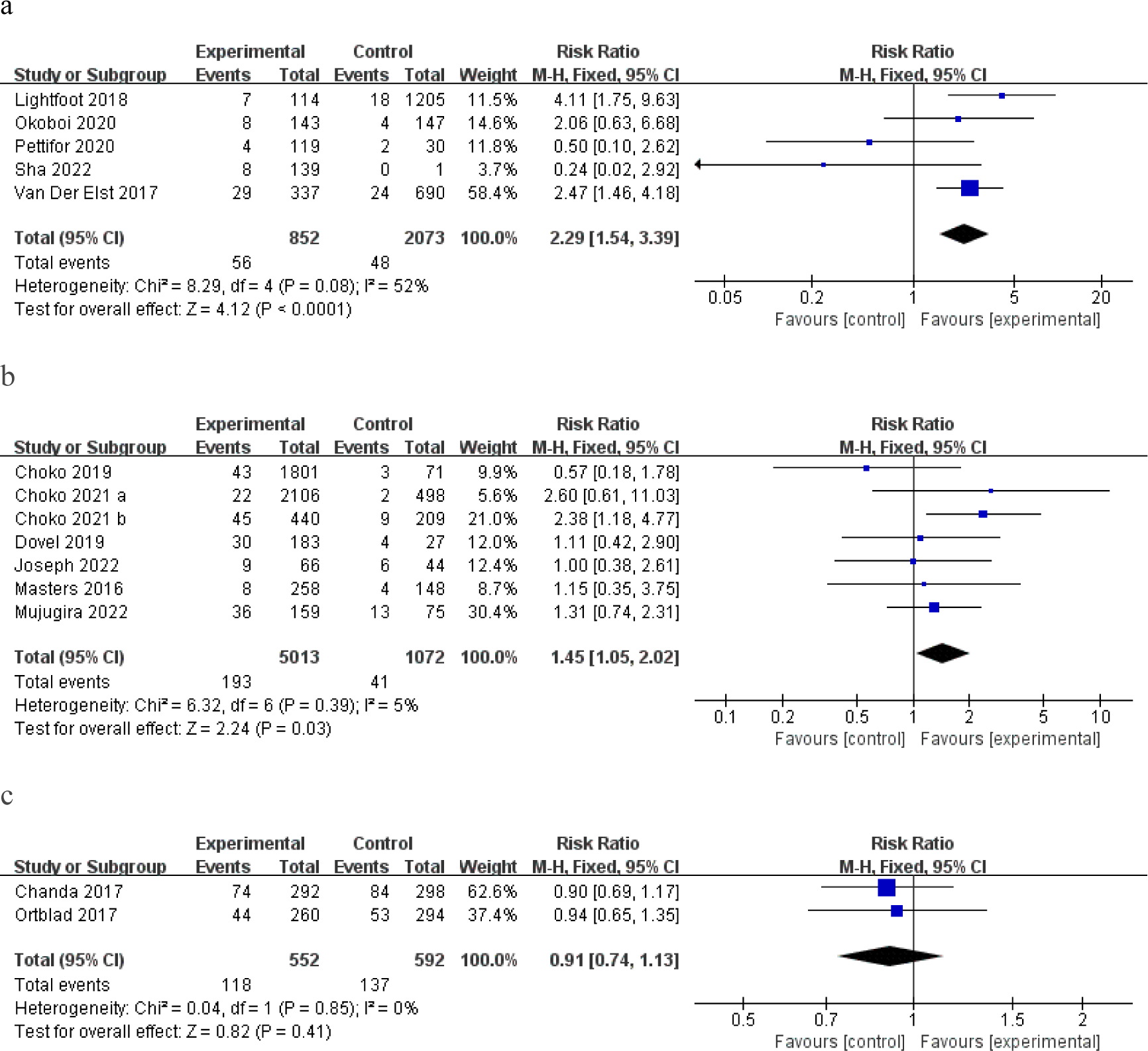
HIV seroconversion by HIVST Distribution Strategy. figure 6(a): Peer distribution in community, figure 6(b): Partner distribution in community, figure 6(c): Peer educator distribution in community.

### Linkage to ART or HIV Care among People Living with HIV

Four direct comparisons contributed to the meta-analysis of the impact of distribution strategies on linkage to ART or HIV care among people living with HIV (Supplementary Table 4). All distribution strategies, whether peer distribution, partner distribution, or peer educator distribution, has comparable linkage to care as facility-based testing (Combined three distributions: RR 0.91, 95% CI 0. 80-1.03, I2 2%). A cohort study directly demonstrated the impact of peer distribution strategies for delivering HIVST kits on linkage: although comparable to the impact of facility-based testing, men who received confirmatory testing after receiving HIVST tended to receive immediate treatment with ART [48].

## Discussion

Our systematic review and meta-analysis examined HIVST distribution strategies using social network strategies. This study extends the existing literature by using network meta-analysis to summarize the social network strategies for HIVST, pooing the effectiveness of augmenting testing rates among at-risk populations, and including linkage to care data. Our results indicated that all HIVST distribution strategies effectively increased HIV testing uptake and testing yield compared to traditional HIV testing performed by HCWs in health facilities, whether peer, partner, or peer educator distribution strategies.

Network-based HIVST distribution could draw on key population assets and community strengths. The results of our study suggest that social network strategies can effectively promote HIV and increase the uptake of HIV testing among at-risk populations. Consistent with past research, HIVST surmounted multifaceted structural impediments besetting HIV testing services, while social network strategies further enhanced coverage on this basis [5, 19, 56]. Social network strategies focus first on risks, recruiting the first at-risk group or people living with HIV for testing, training, and education [14]. Furthermore, social network strategies not only reach these potentially marginalized at-risk populations hidden from current HIV testing practices but also encourage conversations within the social networks about risky behaviors and HIV testing-related information. Based on this asset-based framework, the recruited initial peers are considered sources of wisdom, insight, and strength. Social networks provide greater access to a wide range of risks, health information, and practices, and information can be passed easily and frequently between individuals [57, 58]. In addition, members of the same social network often have similar norms, attitudes, and HIV risk behaviors, and social networks can influence risk and health behaviors through a variety of psychosocial mechanisms and linkage characteristics, such as frequency of contact, duration of contact, social influence, social norms, and social support [59]. Social networks can be used to promote HIVST and to follow up with self-testers. The social influence of peer educators can also be used to disseminate HIV risk information and attempt to change social norms to increase HIV prevention and testing behaviors [36–38]. Consequently, the systematic integration of social network strategies and leverage of community strength for promulgating HIVST warrants earnest endorsement to attain comprehensive and recurrent HIV testing.

Our network meta-analysis showed that HIV testing uptake was highest for distributing HIVST kits through peers and partners. Based on pre-existing contacts, peers and partners spontaneously distribute kits within their social networks [14, 32]. In contrast, peer educators were primarily randomly assigned to groups of recruited participants and then established connections for behavioral interventions [37, 41]. This fact suggests that the distribution strategy of peer educators should be further optimized by considering the dynamics of relationships within the social network of the target population, combining economic costs and social support [60].

Future refinements of HIVST distribution strategies should account for factors such as motivation, skill proficiency, self-efficacy, social norms, behavioral patterns, and supplementary interventions to augment the effectiveness of the peer educator approach [61].

This systematic review and meta-analysis showed that the social network intervention was associated with increased testing yield. Consistent evidence suggests that social network intervention has proven effective in identifying undiagnosed HIV infection in high-risk networks [62, 63]. Compared to healthcare workers, peers, and partners can interact more interpersonal interactions with members of their social networks and provide them with more authentic empathy, validation, and practical advice, thus providing effective social support for undiagnosed HIV infections in high-risk networks and facilitating case identification [60].

People living with HIV are an important source of infection for the ongoing transmission of HIV, and increasing the testing rate of finding people living with HIV is essential to facilitate linkages to ART or HIV care and prevention services for populations at high risk of HIV. In addition to assisted partner notification services, case identification should be facilitated by expanding service options, especially through social network strategies to convey relevant information [64].

We also found that social network-based HIVST distribution strategies and facility-based testing are comparable in linkage to care. However, social network strategies showed significant improvements in the uptake of testing, which positively impacted linkage. In addition, peer-based interventions, including communication links, social support, and monetary incentives, increased the likelihood of linkage to care [65, 66]. The comparable linkages demonstrated the reliability and potential of the social network strategy in improving HIV care services, not only in HIV testing. Future network studies should further consider and evaluate additional robust interventions to support the linkage between post-HIVST testing and HIV care.

This systematic evaluation and meta-analysis have several limitations. First, 31 of the 33 studies had a high or unclear risk of bias in at least one of the methodological quality assessments. Due to the nature of the intervention and social network strategies to distribute HIVST kits, it was difficult to conduct blinded comparisons of participants assigned to study groups or study allocators. Second, the methods used in the comparison group were not entirely consistent. For example, in the comparison group, either no intervention was provided, or HIVST services were promoted without peer educators or the use of social network strategies to provide general health-related information. We further categorized intervention types to mitigate the heterogeneity and scrutinized measurement heterogeneity before combining the data. Third, the included studies varied widely in terms of intervention duration and time to outcome assessment, leading to potential bias in this meta-analysis.

The discerned findings primordially and compellingly advocate for the progressive amplification of peer and partner distribution strategies, underpinned by empirical evidence that underscores their remarkable efficacy. First, the assets and community power of the key population should be fully utilized and mobilized. Similarly, in its new recommendations on HIV testing, WHO is calling on countries to increase testing coverage by promoting testing through sexual and social networks [8]. Second, more efficient distribution strategies for peer educators should be further explored, considering economic costs and incorporating the dynamics of relationships within the social networks of the target population. Future HIV prevention interventions could be carried out in partnership with the community to identify trusted and knowledgeable peer educators and train them to be most effective [67]. Finally, along with increasing testing for early detection of people living with HIV, emphasis should be placed on increasing access to ART or HIV care among at-risk populations. Peers play an important role in providing support for linkage to care services. Different additional interventions, such as home-initiated ART care [68], conditional monetary incentives [50, 52], and peer educator navigating [29] could be chosen to improve the linkage after HIVST. It is paramount to acknowledge that the potency of social network interventions is contingent on their continual evolution and alignment with evolving paradigms. To harness the true potential of social networks in curbing HIV transmission, it is incumbent to propel dedicated research endeavors and maintain an ongoing regimen of scrutiny and adaptation. These imperative measures are indispensable in not only curbing the onward trajectory of HIV transmission but also in charting a course that fortifies linkage to ART or HIV care subsequent to HIV infection.

## Conclusions

This comprehensive systematic review and meta-analysis stand as a testament to social network strategies’ viability, acceptance, and efficacy in promoting HIVST. The findings therein substantiate the resounding impact of diverse HIVST distribution strategies, universally augmenting the uptake of HIV testing, facilitating the early identification of cases, and effectively linking to HIV care. It is necessary to capitalize on the assets and community strengths of the key population. The ascendancy of interventions at the social network stratum extends beyond mere testing proficiency; they furnish a dynamic platform for dispensing services and instigating shifts in social norms. Consequently, these transformations precipitate salubrious changes in risk and health behaviors that orchestrate a positive ripple effect on HIV outcomes.

## Authors’ contributions

W. T. and J. D. designed the review. C. F., Y. D., and S. H. identified relevant studies and extracted data. Y. X., Y. Z. and H. L. analyzed and interpreted data. All other authors (X. H., D. W., J. T. and W. T.) contributed to data interpretation. S. H. and F. J. wrote the first draft of the article. All authors critically revised the article and approved the final version.

## Data Availability

All data produced in the present work are contained in the manuscript

## Acknowledgements

Funding

This work was supported by the Key Technologies Research and Development Program (2022YFC2304900-4 to WT), National Institute of Health (R34MH119963, 1UG1HD113156-01, 1R25AI170379-01 and R01AI158826 to WT), National Nature Science Foundation of China (81903371 to WT), and CRDF Global (G-202104-67775 to WT). The funders had no role in study design, data collection and analysis, decision to publish, or preparation of the manuscript.

## Supporting Information

Supporting Information file 1: Supplementary Appendix 1. Search terms. Supporting Information file 2: Supplementary materials.

## List of abbreviations

ANC: Antenatal Care Clinic
ART: Antiretroviral Therapy
CI: Confidence interval
CrI: Credible intervals
FSW: Female Sex Workers
HCW: Healthcare Worker
HIV: Human Immunodeficiency Virus
HIVST: HIV Self-testing
HTS: HIV Testing Services
MSM: Men Who Have Sex with Men
PWID: People who Inject Drugs
RR: Relative Risk
RCT: Randomized Controlled Trial
SOC: Standard of Care
SUCRA: Surface under the Cumulative Ranking
WHO: World Health Organization

