## Supplementary Appendix 1 for "Social Network Strategies to Distribute HIV Self-testing Kits: A Global Systematic Review and Network Meta-analysis"

**Table S1 Search terms**

| No. | Terms | Comments |
|---|---|---|
| 1 | (((((((((((((((((((((((((((((((((((((((((((((("HIV")) OR "Acquired Immunodeficiency Syndrome") OR (HIV)) OR (Human Immunodeficiency Virus)) OR (Immunodeficiency Virus, Human)) OR (Immunodeficiency Viruses, Human)) OR (Virus, Human Immunodeficiency)) OR (Viruses, Human Immunodeficiency)) OR (Human Immunodeficiency Viruses)) OR (Human T Cell Lymphotropic Virus Type III)) OR (Human T-Cell Leukemia Virus Type III)) OR (LAV-HTLV-III)) OR (Lymphadenopathy-Associated Virus)) OR (Lymphadenopathy Associated Virus)) OR (Lymphadenopathy-Associated Viruses)) OR (Virus, Lymphadenopathy-Associated)) OR (Viruses, Lymphadenopathy-Associated)) OR (Human T Lymphotropic Virus Type III)) OR (Human T-Lymphotropic Virus Type III)) OR (AIDS Virus)) OR (AIDS Viruses)) OR (Virus, AIDS)) OR (Viruses, AIDS)) OR (Acquired Immune Deficiency Syndrome Virus)) OR (Acquired Immunodeficiency Syndrome Virus)) OR (HTLV-III)) OR (HIV-1)) OR (HIV-2)) OR (HIV1)) OR (HIV2)) OR (Acquired Immunodeficiency Syndrome)) OR (Immunologic Deficiency Syndrome, Acquired)) OR (Acquired Immune Deficiency Syndrome)) OR (Acquired Immuno-Deficiency Syndrome)) OR (Acquired Immuno Deficiency Syndrome)) OR (Acquired Immuno-Deficiency Syndromes)) OR (Immuno-Deficiency Syndrome, Acquired)) OR (Immuno-Deficiency Syndromes, Acquired)) OR (Syndrome, Acquired Immuno-Deficiency)) OR (Syndromes, Acquired Immuno-Deficiency)) OR (Immunodeficiency Syndrome, Acquired)) OR (Acquired Immunodeficiency Syndromes)) OR (Immunodeficiency Syndromes, Acquired)) OR (Syndrome, Acquired Immunodeficiency)) OR (Syndromes, Acquired Immunodeficiency)) OR (AIDS) | HIV/AIDS |

| | | |
|---|---|---|
| 2 | ("Social Networking" OR "Social Network Analysis" OR "Social Networking" OR "Networking, Social" OR "Social networks" OR "network social" OR "Social network" OR "Social Network Analysis" OR "Analyses, Social Network" OR "Analysis, Social Network" OR "Network Analysis, Social" OR "Social Network Analyses" OR "peer" OR "peer influence" OR "peer leader" OR "peer educator" OR "peer mentor" OR "seed" OR "ego" OR "index" OR "index participant" OR "alter" OR "support") | Social network |
| 3 | "hiv self-testing" OR "hiv self-test" OR "HIVST" OR "home test" OR "rapid test" OR "home self test" OR "Self-test" OR "Self-testing" OR "home test" OR "home testing" | HIVST |
| 4 | (#1 AND #2 AN D #3) | |
