## Supplementary materials for "Social Network Strategies to Distribute HIV Self-testing Kits: A Global Systematic Review and Network Meta-analysis"

### S1(a) Table: HIVST distribution strategy details - No comparison group

| Study | Method of identifying seeds | Method of identifying network members | Role of social network | HIVST Delivery Method | Who distributed HIVST | Where HIVST delivered | How HIVST delivered | Testing choice offered | Testing support | Additional intervention components |
|---|---|---|---|---|---|---|---|---|---|---|
| Kitenge 2022 | At PHCs, lay counsellors invited all users to participate. At community-based testing sites, CHWs introduced the study to all individuals visiting these sites. | PRs distributed HIVST kits to their sexual partner, family members or anyone in their social network who was above 18 years old. | Accessing the hard to-reach & Diffusion | Secondary | Lay counsellors or community health workers to peer to social network | Social network | In person | No | Instruction on use | No |
| Kwan 2023 | Seed participants were recruited from various internet platforms, including social media platforms, a web-based forum, and location-based networking apps used by MSM. | Seed MSM participants invited their peers. | Accessing the hard to-reach & Diffusion | Secondary | Web-based platform to participants to social network | Online | Mail | Yes (finger-prick or oral fluid test) | Real-time support, including in-person, video call, and instant messaging support, at kit request. | Monetary incentive (To encourage return, an incentive of HK $20 (US $2.56) was given after manual verification of the result. ) |
| Lippman 2018 | Participants were recruited from all HIV-negative MpMS. | Participants shared the kits with sexual partners and others with whom they felt safe distributing kits | Accessing the hard to-reach & Diffusion | Secondary | HCW to participants to social network | social network | In person | HIVST (oral fluid or blood) | All participants were shown a demonstration on how to use both the oral fluid and fingerstick HIV self-tests. | No |

| Study | Method of identifying seeds | Method of identifying network members | Role of social network | HIVST Delivery Method | Who distributed HIVST | Where HIVST delivered | How HIVST delivered | Testing choice offered | Testing support | Additional intervention components |
|---|---|---|---|---|---|---|---|---|---|---|
| Li S 2021 | 6 key opinion leaders of MSM disseminated an HIVST recruitment advertisement to invite MSMs | IPs shared the HIVST service links on social media with their sexual partners and friends. | Accessing the hard to-reach & Diffusion | Online & mail | HIVST application link for phone app to index to alter | Online | Mail | No | HIV posttest consultation | Monetary incentive (US $2 was paid to participants who uploaded their test results) |
| Matovu 2020 | Community residents selected one peer-leader per social network grouping | Peer-leaders distributed kits to eligible social network members. | Promote HIVST and increase linkage to HIV care among newly diagnosed HIV-positive individuals | Community -based | HCW to Peer-educators to social network | In community | In person | No | Peer-leaders received a training on oral HIV self-testing processes; basic counselling, communication and referral skills; and how to approach social network members at the time of distributing the kits;a practical demonstration of how the HIV self-testing exercise is conducted. | No |
| Nasuuna 2022 | Recruitment of KP peers and pregnant and lactating mothers | HIVST was implemented using peer-to-peer model for KPs and secondary distribution for partners of consenting pregnant and lactating mothers. | Accessing the hard to-reach & Diffusion | Secondary | peer-to-peer/HCW to participants to partner | In community | In person | No | 3-day trainings prior to distribution of HIVST kits. Education on how to use HIVST kits was provided in the local language | Phone call (Mothers and peers were asked to confirm delivery of the kits to recipients through a phone call.) |

| Study | Method of identifying seeds | Method of identifying network members | Role of social network | HIVST Delivery Method | Who distributed HIVST | Where HIVST delivered | How HIVST delivered | Testing choice offered | Testing support | Additional intervention components |
|---|---|---|---|---|---|---|---|---|---|---|
| Nguyen 2019 | KPs and their partners were offered HIVST by peer-educators | Community-led outreach and social networks were used to promote HIVST. | Promote HIVST and follow-up with self-testers | Community -based | HCW to Peer-educators to KP and their partners | In community | In person | Self-testers were given the choice to test with or without assistance. | Assisted HIVST | No |
| Nguyen 2019 | KPs and their partners were offered HIVST by peer-educators | Both in-person and social network methods were used to mobilize key populations to test for HIV and offer HTS to partners of people living with HIV. | mobilize key populations to test for HIV and offer HTS to partners of people living with HIV | Community -based | HCW to Peer-educators to KP and their partners | In community | In person | Yes: lay provider-delivered rapid testing / HIVST | Peer educators were trained to conduct rapid testing, perform and demonstrate self-testing, provide pre-test information and post-test counselling, and deliver aPN. | No |
| Thirumurthy 2016 | HIV-uninfected women aged 18–39 years were recruited at a health facility with antenatal (ANC) and postpartum (PPC) clinics, and a drop-in center for female sex workers (FSW). | Male partners HIV-uninfected women | Increased coverage of HIV testing services for partners and the odds of partners being linked to care or prevention | Secondary | HCW to women with high HIV incidence to their sexual partners | Partners among HIV-uninfected women | In person | No | Instructions on using the OraQuick Rapid HIV 1/2 Test and telephone support | No |

| Study | Method of identifying seeds | Method of identifying network members | Role of social network | HIVST Delivery Method | Who distributed HIVST | Where HIVST delivered | How HIVST delivered | Testing choice offered | Testing support | Additional intervention components |
|---|---|---|---|---|---|---|---|---|---|---|
| Wu D 2021 | MSMs were recruited from a social-media based online system | Index MSMs distributed the kits to other social contacts, including partners or friends | Accessing the hard to-reach & Diffusion | Secondary | HIVST application link for phone app to index to alter | Social network | Mail | No | Instruction on use and HIV post-test consultation | Monetary incentive (US$3 was provided to all participants who completed a questionnaire. ) |
| Zhang J 2021 | Participants were recruited from MSM in the ongoing PrEP pragmatic trial | Participants shared HIVST with their male sexual partners. | Increased coverage of HIV testing services for partners and the odds of partners being linked to care or prevention | Secondary | HCW to participants to partner | partners among MSM | In person | No | A service account on WeChat provided web-based services on the application of extra testing kits, instructions on self-testing, real-time consultation with the staff, uploading of test outcomes, and follow-up questionnaires. | Daily or on-demand PrEP |
| Zishiri 2022 | Antenatal care (ANC) clinic attendees and people or those newly diagnosed with HIV clients were given HIVST kits. | Clinic attendees distributed HIVST kits to their partners. | Increased coverage of HIV testing services for partners and the odds of partners being linked to care or prevention | Secondary | HCW to participants to partner | In community | In person | No | Instructions and demonstration of how to use | No |

### S1(b) Table: HIVST distribution strategy details - Comparison group

| Study | Method of identifying seeds | Method of identifying network members | Randomization method | Role of social network | HIVST Delivery Method | Who distributed HIVST | How HIVST delivered | Testing choice offered | Testing support | Additional intervention components | Comparison |
|---|---|---|---|---|---|---|---|---|---|---|---|
| Chanda 2017 | Recruitment of current or former FSWs as peer educators | Participants were recruited by peer educators. | Peer educator–participant groups were randomized as clusters in a 1:1:1 fashion to 1 of the 3 study arms | Diffusion & Impact on risk and health behaviors | Facility & secondary by peer | HCW to Peer-educators to participants | In person | No | Instructions for use and telephone support | NA | Routine facility-based HIV testing |
| Choko 2019 | Women aged 18 years and older attending an ANC at primary care clinics, whose primary male partner was not known to be on ART were recruited. | Women were given an invitation letter addressed to their male partner informing them of the importance of having an HIV test | We block randomized ANC days (representing clusters of women) to 1 of 6 trial arms (standard of care [SOC] and 5 intervention arms; ratio 1:1:1:1:1:1). | Increased coverage of HIV testing services for men and the odds of men being linked to care or prevention | Secondary | HCW to participants to partners | In person | No | Instructions for use | Monetary incentive ($3, $10, and a phone reminder) | Routine facility-based HIV testing |
| Choko 2021 | Eligible women attending ANC were enrolled into the ANC clinic cohort and people newly diagnosed with HIV during routine clinic HIV testing were enrolled into the index cohort. | Eligible women provided with materials and brief training for their male partner. Index patients delivered materials to all sexual contacts over the past 12 months. | Restricted randomization (blocking) was used to randomize the 27 clusters to three arms in a ratio of 1:1:1, with district and HIV prevalence in ANC as the variables for restriction. | Accessing the hard to-reach & Diffusion | Secondary | HCW to participants to partner | In person | No | Instructions and demonstration of how to use | Monetary incentive | Routine facility-based HIV testing |

| **Study** | **Method of identifying seeds** | **Method of identifying network members** | **Randomization method** | **Role of social network** | **HIVST Delivery Method** | **Who distributed HIVST** | **How HIVST delivered** | **Testing choice offered** | **Testing support** | **Additional intervention components** | **Comparison** |
|---|---|---|---|---|---|---|---|---|---|---|---|
| Dovel 2019 | Individuals living with HIV and on ART were recruited during routine ART clinic visits. | Partners of ART clients | Computer-generated randomization was used to assign clients to either the PRS or HIVST arms in a ratio of 1:2·5, respectively. | Increased coverage of HIV testing services for partners and the odds of partners being linked to care or prevention | Secondary | HCW to participants to partner | In person | No | Instructions and demonstration of how to use | NA | Routine facility-based HIV testing |
| Frye 2021 | Recruitment was conducted via online advertising, face-to face outreach and referrals by study participants. | Participants completed an online contact card and sent a link to a friend. | Participants were randomized as friend pairs in a 1:1 ratio into either the TRUST intervention or the time- and attention-matched control intervention arm using assignments generated by the study data analyst using Sealed Envelope Ltd. | Impact on risk and health behaviors. TRUST was designed to increase uptake of consistent (every three months) HST among YBMSM/TW in New York City. | Direct | HCW to participants | In person or Mail-in | No | Instructions for use and peer-based behavioral intervention | Monetary incentive | Time and attention control arm |
| Gichangi 2018 | The health facility nurse identified women attending first ANC and referred them to the trained study nurse. | Partner distribution in community | Participants were individually randomized into 1 of the 3 study arms. | Increased coverage of HIV testing services for men and the odds of men being linked to care or prevention | Secondary | HCW to participants to partners | In person | No | Instructions for use | Advise on partner negotiation and communication | Standard information card to invite male partners to the clinic for routine or HIV care |

| Study | Method of identifying seeds | Method of identifying network members | Randomization method | Role of social network | HIVST Delivery Method | Who distributed HIVST | How HIVST delivered | Testing choice offered | Testing support | Additional intervention components | Comparison |
|---|---|---|---|---|---|---|---|---|---|---|---|
| Joseph 2022 | Recruitment of WLHIV who were attending HTS, HIV care/treatment services or antenatal care services by the research team and clinic staff | WLHIV`s male partners | Participants were randomized 1:1 using a random number table by participant identifier (ID) | Increased coverage of HIV testing services for men and the odds of men being linked to care or prevention | Secondary | HCW to participants to partner | In person | No | Instructions for use and telephone support | NA | Routine facility-based HIV testing |
| Lightfoot 2018 | The peer recruiters were identified from HIV-related support groups, local gay bars, online social networking and dating apps, community-based organizations, and word of mouth. | The peer recruiters distributed OraQuick oral fluid HIV test kits to friends who they believed were AAMSM or LMSM, age 18–45, and had not tested in the last year. | N/A | Peers, who were most effective at reaching infrequent, and non-testers. | Secondary | HCW to Peers to social network | In person | No | Instructions for use | Monetary incentive(Each peer received $150 after distribution of all 5 HIVST tests. Testers had the alternative option to provide contact information for receipt of cash or another gift card for $25. | Testers in County Testing Program |
| MacGowan 2020 | Persons who clicked the ads on social network, music, and dating websites were directed to research sites to complete eligibility screening. | Distribution HIV self-tests to social network members | Participants were randomly assigned to the self-testing (ST) arm or control arm using a computer-generated stochastic random 1:1 allocation. | Accessing the hard to-reach & Diffusion | Secondary | Website online to index to alter | mail | No | Instruction enhancement, video or study hotline | NA | Website with routine HIV testing information |
| Masters 2016 | Trained research assistants screened and enrolled women seeking ANC or PPC at the three facilities, in a private location away from regular clinic activities. | Partner distribution in community | Participants were randomized in a 1:1 ratio using balanced block randomization (block size 20) to an HIVST group or a comparison group. | Increased coverage of HIV testing services for men and the odds of men being linked to care or prevention | Secondary | HCW to participants to partners | In person | No | Instructions and demonstration of how to use | Advise on partner negotiation and communication | Invitation card for clinic-based HIV testing |

| Study | Method of identifying seeds | Method of identifying network members | Randomization method | Role of social network | HIVST Delivery Method | Who distributed HIVST | How HIVST delivered | Testing choice offered | Testing support | Additional intervention components | Comparison |
|---|---|---|---|---|---|---|---|---|---|---|---|
| Merchant 2018 | Via multiple social media platforms | YMSMs recommended three other 18–24-year-old black, Hispanic, or white YMSM. | Within each racial/ethnic group, we randomly assigned participants into one of three study arms (1:1:1 randomization) using block sizes of six. | Accessing the hard to-reach & Diffusion | Online & mail | Website/ Study app to participants to social network | Mail | No | No | Monetary incentive (Participants received a $10 internet-based gift card for completing the follow-up questionnaire, and up to three $5 gift cards for providing email addresses of other YMSM to be contacted about the study. ) | Routine facility-based HIV testing |
| Mujugira 2022 | PWLHIV attending antenatal care who reported that their partner's HIV status was unknown. | Secondary distribution of HIV self-test kits (HIVST) from HIV-negative pregnant women to their partners. | Eligible participants were randomized 2:1 to HIVST secondary distribution or an invitation for fast-track HIV testing for their partner. | Increased coverage of HIV testing services for men and the odds of men being linked to care or prevention | Secondary | HCW to participants to partner | In person | No | trained in the use and interpretatio n of HIVST | NA | Routine facility-based HIV testing |
| Okoboi 2020 | Fifteen MSM peers who trained in HIVST testing procedures and results interpretation | Peers distributed HIVST kits to MSM in their social and sexual networks. | N/A | Increasing testing coverage among non-testers. | Secondary | HCW to Peers to social network | In person | No | Instructions and demonstrati on of how to use and telephone support | Transport reimbursement and T-shirts, backpacks and umbrellas as tokens of appreciation | Routine facility-based HIV testing |

| Study | Method of identifying seeds | Method of identifying network members | Randomization method | Role of social network | HIVST Delivery Method | Who distributed HIVST | How HIVST delivered | Testing choice offered | Testing support | Additional intervention components | Comparison |
|---|---|---|---|---|---|---|---|---|---|---|---|
| Ortblad 2017 | Recruitment of FSW peer educators | Peer-educators recruitment, research assistants telephone screening, eligibility assessment, and enrollment. | We randomized FSW peer educator groups 1:1:1 to 1 of 3 study arms. | Have good access to other FSWs and are able to engage with FSWs who do not normally utilize the health system & Diffusion & Impact on risk and health behaviors | Facility & secondary by peer | HCW to Peer-educators to participants | In person | No | Instructions for use and telephone support | NA | Routine facility-based HIV testing |
| Pettifor 2020 | Recruitment of young women from Agincourt Health and Social Demographic Surveillance System (AHDSS) | Young women offered test kits/invitations to peers and sexual partners. | Young women ages 18-26 were randomized using block randomization with a 1:1 allocation to either | Secondary distribution of HIV self-test kits is one way to reach populations who test less frequently and hard-to-reach groups. | Secondary | HCW to index to alter | In person | Yes (choice of either a clinic-based HCT invitation or oral HIV Self-Testing (HIVST) kits.) | Instructions for use, include pre- and post-test counseling information and a “frequently asked questions” document on HIV self-testing | NA | Routine facility-based HIV testing |
| Sha 2022 | Community volunteers or public health workers invited men who visited the three clinics for HIV testing. | Indexes distributed HIV/syphilis dual self-testing kits to people in their social networks. | The two arms were implemented one at a time to compare the intended outcomes in similar catchment areas without having people choose between the two. | Accessing the hard to-reach & Diffusion | Secondary | HCW to index to alter | In person | No | Instructions for use | Monetary incentive (Indexes received $3 after completing the baseline survey and $5 for a follow-up survey. Alters and corresponding indexes both received an additional $3 when alters uploaded their test results.) | Testing card referral (facility-based test) |

| Study | Method of identifying seeds | Method of identifying network members | Randomization method | Role of social network | HIVST Delivery Method | Who distributed HIVST | How HIVST delivered | Testing choice offered | Testing support | Additional intervention components | Comparison |
|---|---|---|---|---|---|---|---|---|---|---|---|
| Shahmanesh 2021 | Recruitment of 24 pairs of peer navigators through local municipal and traditional leaders | Peer navigators recruited young people in community settings near schools and homes. | The final groupings of peer navigators into three arms were completed using statistical software, into three groups of 8 pairs and three floating peer navigators (A, B and C). | Diffusion & Impact on risk and health behaviors | Facility & secondary by peer | HCW to Peer-educators to participants | In person | No | In all three arms, peer navigators promoted sexual health and the benefits of HIV testing PrEP and ART. In both intervention arms, they also demonstrated how to use the HIVST kit. | Monetary incentive | Routine facility-based HIV testing |
| Young 2013 | 18 peer leaders were recruited from community organizations serving African American and Latino MSM. | Participants were recruited from ads on Internet and social networking sites | Participants were randomly and blindly assigned to 1 of 2 intervention or control groups and then randomly assigned to 2 peer leaders within that group. | Diffusion & Impact on risk and health behaviors. The community peer leader model is designed to increase HIV prevention and testing behaviors by changing social norms. | Direct | HCW to Peer-educators to participants | Mail-in | No | Instructions for use | NA | General health information |

| Study | Method of identifying seeds | Method of identifying network members | Randomization method | Role of social network | HIVST Delivery Method | Who distributed HIVST | How HIVST delivered | Testing choice offered | Testing support | Additional intervention components | Comparison |
|---|---|---|---|---|---|---|---|---|---|---|---|
| Young 2022 | 79 peer leaders were recruited with help from community organizations serving Latinx and African American MSM. | From online advertisements on Facebook, Craigslist, and other websites/applications; community physical venues frequented by Latinx and African American MSM; and from direct referrals from study participants. | Using a random number generator with participants blinded to assignment and unable to request group or condition assignment. | Diffusion & Impact on risk and health behaviors. The community peer leader model is designed to increase HIV prevention and testing behaviors by changing social norms. | Direct | HCW to Peer-educators to participants | Mail-in | No | Instructions for use and communicate in the online community, by sending messages, chats, and wall posts | NA | Non-peer-led HIVST |
| Van Der Elst 2017 | Mobilizations were done through GBMSM peers from a local GBMSM-led community group | GBMSM peers mobilized in community | NA | Accessing the hard to-reach & Diffusion | Secondary | HCW to Peers to social network | In person | No | Close supervision and daily feedback | Participants asked to report for confirmatory HTC at the clinic | Routine facility-based HIV testing |
| Zhou 2022 | Volunteers of Xutong enrolled participants by advertising the recruitment and trial introduction in the WeChat public platform. | Index participants distributed HIVST kits to members within their social networks. | Eligible participants were randomly assigned to one of the 3 arms individually and independently by a computer-generated program electronically. | Accessing the hard to-reach & Diffusion | Secondary | HCW to index to alter | Mail-in | No | Instructions for use | Monetary incentive (Index participants in the 2 intervention groups could receive a fixed incentive ($3 USD) online for the verified test result uploaded to the digital platform by each unique alter.) | Standard secondary distribution |

### S2 Table: Outcomes of the included studies

| Study | Study arm | HIV testing uptake | | | HIV seroconversion | | | Linkage to ART or HIV care among HIV positive | | | |
|---|---|---|---|---|---|---|---|---|---|---|---|
| | | Time point | Offered HIVST | Tested for HIVST | Time point | All positive | Tested for HIVST1 | Time point | ART initiation | Linked to HIV care | HIV positive |
| Chanda 2017 | Standard of care | 2016.9-2017.2 | 1mom: 296, 4mon: 301 | 1mom: 262, 4mon: 226 | 2016.9-2017.2 | 1mom: 59, 4mon: 84 | 1mom: 288, 4mon: 298 | 2016.9-2017.2 | 1mom: 27, 4mon: 54 | 1mom: 44, 4mon: 72 | 1mom: 59, 4mon: 84 |
| Chanda 2017 | Delivery | 2016.9-2017.2 | 1mom: 296, 4mon: 295 | 1mom: 280, 4mon: 248 | 2016.9-2017.2 | 1mom: 49, 4mon: 74 | 1mom: 294, 4mon: 292 | 2016.9-2017.2 | 1mom: 11, 4mon: 35 | 1mom: 25, 4mon: 53 | 1mom: 49, 4mon: 74 |
| Chanda 2017 | Coupon | 2016.9-2017.2 | 1mom: 294, 4mon: 302 | 1mom: 248, 4mon: 241 | 2016.9-2017.2 | 1mom: 36, 4mon: 77 | 1mom: 291, 4mon: 300 | 2016.9-2017.2 | 1mom: 9, 4mon: 44 | 1mom: 19, 4mon: 59 | 1mom: 36, 4mon: 77 |
| Choko 2019 | Standard of care | 28 days | 408 | 71 | 28 days | 3 | 71 | 28 days | 3 | - | 3 |
| Choko 2019 | Partner distribution in community | 28 days | 1941 | 1801 | 28 days | 43 | 1801 | 28 days | 39 | - | 43 |
| Choko 2021 | Standard of care | 28 days | ANC:1447 index:234 | ANC:498 index:209 | 28 days | index:9 ANC:2 | index:209 | - | ANC:2 | - | index:9 ANC:2 |
| Choko 2021 | HIVST | 28 days | ANC:1465 index:169 | ANC:1106 index:155 | 28 days | index:13 ANC:0 | index:155 | - | ANC:0 | - | index:13 ANC:0 |
| Choko 2021 | HIVST plus | 28 days | ANC:1632 index:305 | ANC:1000 index:285 | 28 days | index:32 ANC:22 | index:285 | - | ANC:22 | - | index:32 ANC:22 |
| Dovel 2019 | Standard of care | 2018.3-2018.6 | 107 | 27 | - | 4 | 27 | 12 months | 3 | - | 4 |
| Dovel 2019 | Partner distribution in community | 2018.3-2018.6 | 258 | 183 | - | 30 | 183 | 12 months | 14 | - | 30 |
| Frye 2021 | Standard of care | 2016.7-2019.1 | 197 | 3 months: 83 6 months: 83 | - | - | - | - | - | - | - |
| Frye 2021 | Peer-based behavioral intervention | 2016.7-2019.1 | 179 | 3 months: 102 6 months: 97 | - | - | - | - | - | - | - |
| Gichangi 2018 | Standard of care | 3 months | 938 | 239 | - | - | - | - | - | - | - |

| | | | | | | | | | | | |
|---|---|---|---|---|---|---|---|---|---|---|---|
| Gichangi 2018 | Partner distribution in community | 3 months | 472 | 322 | - | - | - | - | - | - | - |
| Joseph 2022 | Standard of care | 3 months | 85 | 44 | 3 months | 6 | 44 | 3 months | 6 | - | 6 |
| Joseph 2022 | Partner distribution in community | 3 months | 91 | 66 | 3 months | 9 | 66 | 3 months | 6 | - | 9 |
| Lightfoot 2018 | Self-Testers | 2016.1-2017.3 | 165 | 114 | 2016.1-2017.3 | 7 | 114 | - | - | - | 7 |
| Lightfoot 2018 | Testers in County Testing Program | 2016.1-2017.3 | 3483 | 1205 | 2016.1-2017.3 | 18 | 1205 | - | - | - | 18 |
| MacGowan 2020 | Standard of care | 2015.3-2016.11 | - | - | 2015.3-2016.11 | - | - | - | - | - | - |
| MacGowan 2020 | Peer community distribution HIVST | 2015.3-2016.11 | 2864 | 2301 | 2015.3-2016.11 | 34 | 2301 | - | - | 26 | 36 |
| Masters 2016 | Standard of care | 3 months | 286 | 148 | 3 months | 4 | 148 | - | - | 3 | 4 |
| Masters 2016 | Partner distribution in community | 3 months | 284 | 258 | 3 months | 8 | 258 | - | - | 2 | 8 |
| Merchant 2018 | Standard of care | 3 months | 71 | 10 | - | - | - | - | - | - | - |
| Merchant 2018 | mail-testing | 3 months | 57 | 14 | - | - | - | - | - | - | - |
| Merchant 2018 | Online & mail distribution | 3 months | 82 | 17 | - | - | - | - | - | - | - |
| Mujugira 2022 | Standard of care | 12 months | 161 | 75 | 12 months | 13 | 75 | 12 months | 10 | - | 13 |
| Mujugira 2022 | Partner distribution in community | 12 months | 328 | 159 | 12 months | 36 | 159 | 12 months | 25 | - | 36 |
| Okoboi 2020 | Standard of care | 2018.6-2018.8 | - | 147 | 2018.1-2018.3 | 4 | 147 | - | - | - | 4 |
| Okoboi 2020 | Peer community distribution | 2018.6-2018.8 | 150 | 143 | 2018.6-2018.8 | 8 | 143 | - | 8 | 8 | 8 |
| Ortblad 2017 | Peer community distribution HIVST | 2016.10-2017.3 | 1mom: 289, 4mon: 262 | 1mom: 275, 4mon: 261 | 2016.10-2017.3 | 1mom: 39, 4mon: 44 | 1mom: 287, 4mon: 260 | 2016.10-2017.3 | 1mom: 13, 4mon: 19 | 1mom: 17, 4mon: 27 | 1mom: 39, 4mon: 44 |
| Ortblad 2017 | HCW at health facility | 2016.10-2017.3 | 1mom: 321, 4mon: 297 | 1mom: 258, 4mon: 288 | 2016.10-2017.3 | 1mom: 54, 4mon: 80 | 1mom: 312, 4mon: 289 | 2016.10-2017.3 | 1mom: 10, 4mon: 27 | 1mom: 13, 4mon: 37 | 1mom: 54, 4mon: 80 |
| Ortblad 2017 | Standard-of-care | 2016.10-2017.3 | 1mom: 316, 4mon: 302 | 1mom: 226, 4mon: 263 | 2016.10-2017.3 | 1mom: 39, 4mon: 53 | 1mom: 301, 4mon: 294 | 2016.10-2017.3 | 1mom: 13, 4mon: 24 | 1mom: 25, 4mon: 37 | 1mom: 39, 4mon: 53 |
| Pettifor 2020 | Peer community distribution | 3 months | 373 | 119 | 3 months | 4 | 119 | - | - | - | 4 |

| | | | | | | | | | | | |
|---|---|---|---|---|---|---|---|---|---|---|---|
| Pettifor 2020 | Standard of care | 9 months | 406 | 114 | 9 months | 8 | 114 | - | - | - | 8 |
| Pettifor 2020 | Peer community distribution | 9 months | 701 | 393 | 9 months | 14 | 393 | - | - | - | 14 |
| Sha 2022 | Secondary distribution | 2019.5-2020.1 | 179 | 139 | 2019.5-2020.1 | 8 | 139 | - | - | - | 8 |
| Sha 2022 | Testing card referral | 2019.10-2020.1 | 26 | 1 | 2019.10-2020.1 | 0 | 1 | - | - | - | 0 |
| Shahmanesh 2021 | Standard of care | 2019.3-2019.9 | 1098 | - | - | - | - | - | - | 111 | - |
| Shahmanesh 2021 | Direct HIVST distribution | 2019.3-2019.9 | 1480 | - | - | - | - | - | - | 111 | - |
| Shahmanesh 2021 | Peer community distribution | 2019.3-2019.9 | 1585 | - | - | - | - | - | - | 50 | - |
| Young 2013 | Standard of care | 2011.3-2011.6 | 11 | 2 | - | - | - | - | - | - | - |
| Young 2013 | Peer-leaders distribution in community | 2011.3-2011.6 | 25 | 9 | - | - | - | - | - | - | - |
| Young 2022 | Standard of care | 2017.2-2021.1 | 450 | 102 | - | - | - | - | - | - | - |
| Young 2022 | Peer-leaders distribution in community | 2017.2-2021.1 | 450 | 130 | - | - | - | - | - | - | - |
| Van Der Elst 2017 | Standard of care | - | - | - | 2015.7-2015.12 | 24 | 690 | 1 Day(s) | 20/24 | - | 24 |
| Van Der Elst 2017 | Peer community distribution HIVST | - | - | - | 2016.3-2016.6 | 29 | 337 | 14 Day(s) | 24/29 | - | 29 |
| Zhou 2022 | control | 2019.10-2020.12 | 65 | 58 | - | 6 | 58 | - | - | - | 6 |
| Zhou 2022 | SD-M | 2019.10-2020.12 | 107 | 101 | - | 4 | 101 | - | - | - | 4 |
| Zhou 2022 | SD-M-PR | 2019.10-2020.12 | 187 | 185 | - | 5 | 185 | - | - | - | 5 |

HIV testing uptake among all randomized or enrolled; HIV positivity among HIV tested; Linkage to ART or HIV care among HIV positive.

### S3(a)Table: Network meta-analysis relative effects (league) table of HIVST distribution strategies - HIV testing uptake

| | **Facility-based testing** | **Peer-community** | **Partner-community** | **Peer educator-community** |
|---|---|---|---|---|
| **Facility-based testing** | **Facility-based testing** | 2.59 (1.41, 5.20) | 1.95 (1.29, 2.95) | 1.22 (0.68, 2.18) |
| **Peer-community** | 0.38 (0.19, 0.71) | **Peer-community** | 0.75 (0.33, 1.56) | 0.47 (0.19, 1.07) |
| **Partner-community** | 0.51 (0.34, 0.77) | 1.33 (0.64, 3.00) | **Partner-community** | 0.63 (0.30, 1.27) |
| **Peer educator-community** | 0.82 (0.46, 1.47) | 2.14 (0.93, 5.30) | 1.60 (0.79, 3.28) | **Peer educator-community** |

### S3(b)Table: Network meta-analysis ranking probabilities of HIVST distribution strategies - HIV testing uptake

| Testing and distribution strategy | Probability of ranking 1 | Probability of ranking 2 | Probability of ranking 3 | Probability of ranking 4 |
|---|---|---|---|---|
| **Facility-based testing** | 0.00 | 0.00 | 0.24 | 0.76 |
| **Peer-community** | 0.79 | 0.19 | 0.02 | 0.00 |
| **Partner-community** | 0.20 | 0.72 | 0.08 | 0.00 |
| **Peer educator-community** | 0.00 | 0.09 | 0.66 | 0.23 |

### S4 Table: Linkage to ART or Any Care Among people living with HIV by Distribution Strategy, Study Design and Population Subgroup

| Strategy | Design | Population Type | Pooled Risk Ratio | Studies |
|---|---|---|---|---|
| Peer-community | Cohort | MSM | 0.99 [0.78, 1.27] | Van Der Elst 2017 |
| Partner-community | RCT | Male partners of ANC clients | 0.83 [0.60, 1.16] | Choko 2019, Masters 2016, Choko 2021 a |
| | RCT | Partners of HIV positive | 0.79 [0.60, 1.04] | Joseph 2022, Mujugira 2022, Dovel 2019 |
| Peer educators-community | RCT | FSW | 0.80 [0.63, 1.02] | Chanda 2017, Ortblad 2017 |

### S5 Table: Risk of bias for included studies - Observational studies

| Study | Representativeness of exposed cohort | Selection of non-exposed cohort | Ascertainment of exposure | Demonstration that outcome of interest was not present at start of study | Cohorts comparable | Assessment of outcome | Length of follow-up | Loss to follow-up rate | Overall Quality |
|---|---|---|---|---|---|---|---|---|---|
| Kwan 2023 | * | - | * | * | - | * | * | * | Good |
| Kitenge 2022 | * | * | * | * | - | * | * | - | Good |
| Lightfoot 2018 | * | - | * | - | * | * | - | * | Fair |
| Lippman 2018 | * | * | * | * | * | - | * | - | Good |
| Li S 2021 | * | - | * | * | - | - | * | - | Poor |
| Matovu 2020 | * | - | * | * | - | - | * | * | Fair |
| Nasuuna 2022 | * | - | * | * | - | - | * | * | Fair |
| Nguyen 2019 1 | * | * | - | - | - | * | - | - | Poor |
| Nguyen 2019 2 | * | * | * | - | - | * | - | - | Poor |
| Okoboi 2020 | * | * | * | * | - | * | * | - | Good |
| Thirumurthy 2016 | * | * | * | * | - | * | * | * | Good |
| Van Der Elst 2017 | * | * | * | - | - | - | * | * | Fair |
| Wu D 2021 | * | - | * | * | - | * | * | * | Good |
| Zhang J 2021 | * | - | * | * | - | - | * | * | Fair |
| Zishiri 2022 | * | - | * | * | - | - | * | * | Fair |

### S6 Table: Risk of bias for included studies – RCTs and a quasi-experimental study

| Study | Random sequence generation (selection bias) | Allocation concealment (selection bias) | Blinding of participants and personnel (performance bias) | Blinding of outcome assessment (detection bias) | Incomplete outcome data (attrition bias) | Selective reporting (reporting bias) | Other bias |
|---|---|---|---|---|---|---|---|
| Chanda 2017 | Low | Low | Low | Low | Low | Low | Low |
| Choko 2019 | Low | Low | High | High | Low | Low | Low |
| Choko 2021 | Low | Low | High | Low | Low | Low | Low |
| Dovel 2019 | Unclear | Unclear | High | High | Low | Low | Unclear |
| Frye 2021 | Low | Low | High | High | Low | Low | Low |
| Gichangi 2018 | Low | Low | High | High | Low | Low | Low |
| Joseph 2022 | Low | Low | High | Unclear | Low | Low | Low |
| MacGowan 2020 | Low | Low | High | High | High | Low | Unclear |
| Masters 2016 | Low | Low | High | High | Low | Low | Low |
| Merchant 2018 | Low | Unclear | High | High | Unclear | Low | Low |
| Mujugira 2022 | Low | Unclear | High | High | Unclear | Low | Unclear |
| Ortblad 2017 | Low | Low | Low | Low | Low | Low | Low |
| Pettifor 2020 | Low | Low | High | High | Low | Low | Low |
| Sha 2022 | High | High | High | Unclear | High | Low | Low |
| Shahmanesh 2021 | Low | Low | High | Low | Unclear | Low | Low |
| Young 2022 | Low | Low | High | Unclear | Low | Low | Low |
| Young 2013 | Low | Unclear | High | High | Unclear | Low | Low |
| Zhou 2022 | Low | Low | High | Low | Low | Low | Low |

### S1 Figure: Risk of bias for included studies – RCTs and a quasi-experimental study

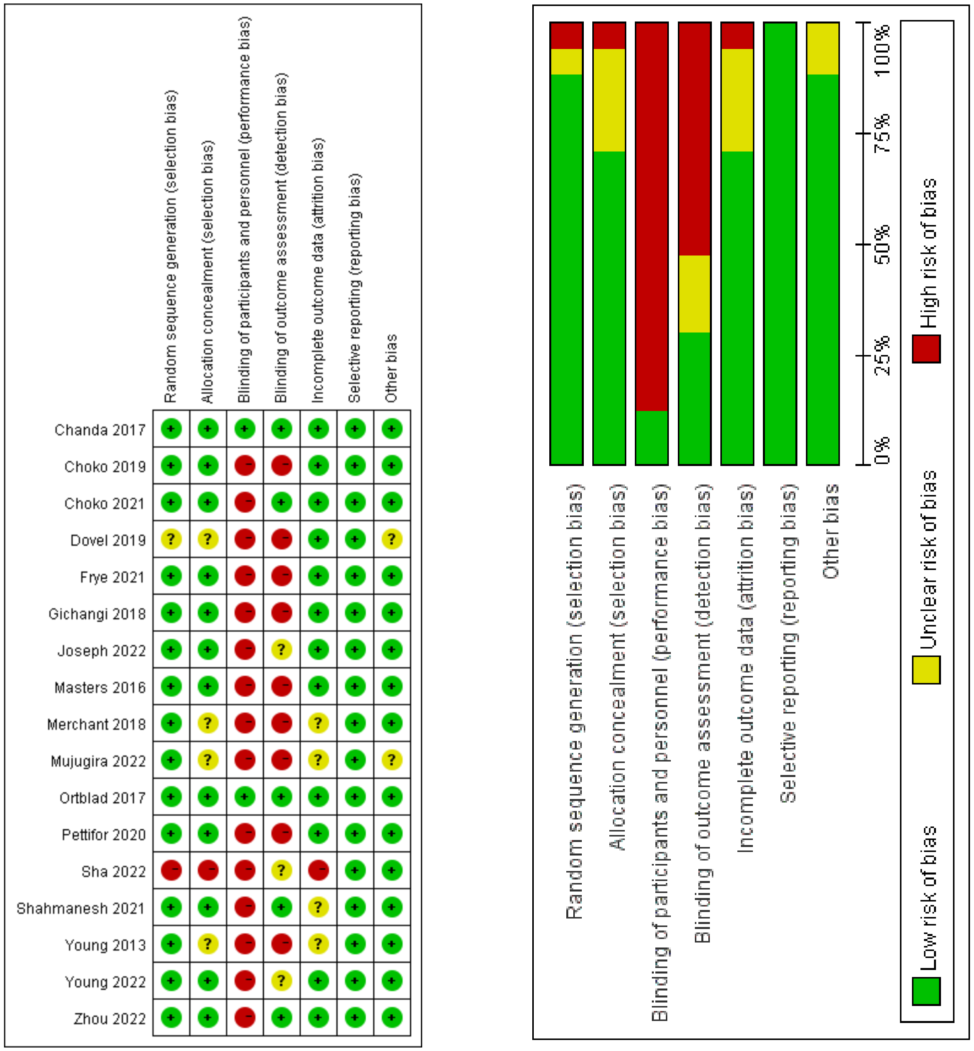

Figure S1. Cochrane risk of bias quality assessment for included studies: RCTs
